# Immunoglobulin E as a biomarker for the overlap of atopic asthma and chronic obstructive pulmonary disease

**DOI:** 10.1101/19004333

**Authors:** Craig P. Hersh, Soumya Zacharia, Ram Prakash Arivu Chelvan, Lystra P. Hayden, Ali Mirtar, Sara Zarei, Nirupama Putcha, the COPDGene Investigators

**Author notes:** Correspondence: Craig P. Hersh, MD, MPH, Channing Division of Network Medicine, Brigham and Women’s Hospital, 181 Longwood Age, Boston, MA 02115. Funding: NIH grants R01HL130512, R01HL125583, K23HL123594, P01HL132825, U01HL089856, and U01HL089897 and a grant from Novartis. The COPDGene study (NCT00608764) is also supported by the COPD Foundation through contributions made to an Industry Advisory Committee comprised of AstraZeneca, Boehringer-Ingelheim, GlaxoSmithKline, Novartis, and Sunovion.

## Abstract

Asthma-COPD overlap (ACO) is a common clinical syndrome, yet there is no single objective definition. We hypothesized that Immunoglobulin E measurements could be used to refine the definition of ACO. In baseline plasma samples from 2870 subjects in the COPDGene Study, we measured total IgE levels and specific IgE levels to six common allergens. Compared to usual COPD, subjects with ACO had higher total IgE levels (median 67.0 vs 42.2 IU/ml) and more frequently had at least one positive specific IgE (43.5 vs 24.5%). We previously used a strict definition of ACO in subjects with COPD, based on self-report of a doctor’s diagnosis of asthma before the age of 40. This strict ACO definition was refined by the presence of atopy, determined by total IgE >100 IU/ml or at least one positive specific IgE, as was a broader definition of ACO based on any asthma history. Subjects will all three ACO definitions were younger (mean age 60.0-61.3), were more commonly African American (36.8-44.2%), had a higher exacerbation frequency (1.0-1.2 in the past year), and had more airway wall thickening on quantitative analysis of chest CT scans. Among subjects with clinical ACO, 37-46% did not have atopy; these subjects had more emphysema on chest CT scan. Based on associations with exacerbations and CT airway disease, IgE did not clearly improve the clinical definition of ACO. However, IgE measurements could be used to subdivide subjects with atopic and non-atopic ACO, who might have different biologic mechanisms and potential treatments.

---

Many patients with chronic obstructive pulmonary disease (COPD) also carry a diagnosis of asthma, termed asthma-COPD overlap (ACO).^1^ ACO has been associated with increased symptoms, increased exacerbations, and greater healthcare utilization. Given the high prevalence and clinical relevance, multiple groups have proposed consensus definitions.^2-4^ The Global Initiative for Asthma (GINA) and the Global Initiative for Obstructive Lung Disease (GOLD) developed a document that uses lung function and clinical factors to determine if an individual patient with airways disease has features more consistent with asthma, COPD or the overlap.^5^ However, many of these factors are subjective and may be difficult to apply in epidemiologic studies or clinical trials.

We have previously reported on ACO within the COPDGene Study, a large observational study of smokers with and without COPD.^6,7^ We defined ACO based on smoking history of at least 10 pack-years, airflow obstruction on spirometry (post-bronchodilator FEV_1_/FVC<0.7 and FEV_1_<80% predicted) and self-report of a physician diagnosis of asthma before the age of 40. Compared to usual COPD, subjects with ACO were younger and were more commonly female and African American. They have a greater number of COPD exacerbations and less emphysema and more airway disease on quantitative analysis of chest CT scans.

Asthma, especially asthma with onset in childhood, has a strong atopic predisposition. The Dutch Hypothesis, put forward in 1961, proposes that there are common host factors for asthma and COPD, including airway hyperresponsiveness and atopy^8^; disease manifestations also depend on external factors, such as exposures. Cigarette smoking and atopy likely increase the risk for COPD development.^9^ COPD subjects with allergic sensitization have been shown to have increased respiratory symptoms and exacerbation rates,^10^ which is similar to our findings in ACO.^6,7^ The finding of a bronchial epithelial type 2 inflammatory gene expression signature in some COPD subjects suggests shared mechanisms with asthma.^11^ However, since bronchoscopy is not part of the standard evaluation in COPD or asthma, a bronchial epithelial gene expression is unlikely to be a clinically-applicable biomarker for ACO.

Therefore, we hypothesized that IgE could be used as a biomarker to refine our previous definition of ACO, which was based on clinical history and spirometry. To test that hypothesis, we measured total and specific IgE levels in a subset of the COPDGene Study, examining clinical and imaging features of subjects with ACO and elevated IgE and/or allergic sensitization.

## Methods

### Study Subjects

The Genetic Epidemiology of COPD Study (COPDGene) is an observational study conducted at 21 clinical centers across the U.S.^12^ COPDGene enrolled non-Hispanic white and African American subjects with a smoking history of at least 10 pack-years. Subjects with previous lung resection and other significant lung diseases, except for asthma, were excluded. The baseline study visit included questionnaires on demographics, medical history, and disease related quality of life, using the St. George’s Respiratory Questionnaire.^13^ Subjects underwent spirometry before and after inhaled albuterol. Exercise capacity was measured by 6-minute walk test. Inspiratory and expiratory chest CT scans with quantitative image analysis were used to assess emphysema (low attenuation areas at -950 HU), gas trapping (-856HU on expiratory scan), and airway wall thickening (wall area percent of segmental airways).^14^ Blood was drawn for genetic analysis and measurement of biomarkers. In the longitudinal follow-up program,^15^ subjects were asked to complete a telephone or web-based questionnaire every six months, assessing incident comorbidities and COPD exacerbations. Exacerbations were defined by the use of antibiotics and/or systemic steroids for a chest illness^16^; severe exacerbations led to an emergency department visit or hospitalization.

All subjects provided written informed consent. The COPDGene Study was approved by the Institutional Review Boards at all participating centers.

### Immunoglobulin E measurements

We selected 2874 COPDGene subjects in four groups defined by the presence or absence of COPD (FEV_1_/FVC<0.7 and FEV_1_<80% predicted, corresponding to GOLD spirometry grades 2-4) and self-reported asthma: asthma, COPD, ACO and controls. Control subjects had normal spirometry (FEV_1_/FVC ≥ 0.7 and FEV_1_ ≥ 80% predicted) and no history of asthma. Plasma samples were send to Phadia Immunology Reference Laboratory (Portage, Michigan) for measurements of total IgE levels and six specific IgE levels using Immunocap assays: cat dander, dog dander, dust mite (D. farinae and D. pteronyssinus), German cockroach and mold mix. For the specific IgE assays, we selected indoor aeroallergens based on a previous COPD study.^10^ To express total IgE as a quantitative outcome, values less than the lower limit of detection (<2 IU/ml) were set as half the lower limit (1 IU/ml) and values greater than assay (>5000 IU/ml) were set at the upper limit of detection (5000 IU/ml). We examined two thresholds for elevated total IgE levels. We used the common clinical cutoff of 100 IU/ml as well as a threshold of 30 IU/ml based on the prescribing information for omalizumab,^17^ a monoclonal antibody against IgE which is indicated for asthma treatment. For five of the specific IgE assays, we defined positive sensitization by a value >0.35 IU/ml. Mold mix was already reported by the lab as positive or negative. For subsequent analyses, atopy was defined by a total IgE level >100 IU/ml or a positive specific IgE to at least one allergen.

### Statistical analysis

In subjects with COPD, we defined “broad” ACO by a self-report of asthma and “strict” ACO by self-report of a doctor’s diagnosis of asthma before the age of 40, as per our previous studies.^6,7^ These groups were further subdivided by the presence of atopy, defined by IgE levels as above. Clinical and imaging features between groups were compared using t-tests or Chi-square tests, as appropriate. The frequency of exacerbations and the outcome of a severe exacerbation were analyzed using linear and logistic regression, respectively, adjusted for age, sex, race, smoking history, and FEV_1_ percent predicted. Analysis of wall area percent of segmental airways used linear regression, adjusted for age, sex, race, current smoking, body mass index, and chest CT scanner model.

## Results

### Study Subjects and IgE measurements

Of the 2874 subjects selected, four subjects were excluded due to spirometry values that did not meet the definitions of COPD or control. Table 1 shows characteristics of study subjects based on COPD and self-report of asthma. Compared to usual COPD, subjects with the broad definition of ACO were younger and more commonly female and African American. Despite fewer pack-years of smoking history, they had lower FEV_1_ and more exacerbations. On quantitative chest CT scan analysis, they had less emphysema and more airway wall thickening. Subjects with ACO had higher total IgE levels than usual COPD, as did subjects with asthma alone compared to controls without airflow obstruction (Table 1). Over 2/3 of ACO subjects had total IgE >30 IU/ml and nearly 40% had total IgE > 100 IU/ml. In ACO, the prevalence of sensitization to the six allergens ranged from 14 to 24%, and over 40% of subjects were sensitized to at least one allergen (Table 1, Supplemental Table 1). These frequencies were higher than usual COPD, but slightly lower than asthma without COPD.

**Table 1:** Study subjects and IgE results. Mean (SD) or N (%) are shown.

|  | Control<br>(no COPD, no asthma) | Asthma<br>(asthma, no COPD) | Usual COPD<br>(COPD, no asthma) | Broad definition ACO<br>(COPD and self-reported asthma) |
| --- | --- | --- | --- | --- |
| N | 598 | 541 | 899<sup>c</sup> | 832<sup>c</sup> |
| Age | 57.2 (8.5) | 55.2 (8.1)<sup>a</sup> | 63.9 (8.5) | 61.9 (8.6)<sup>b</sup> |
| Male gender | 321 (53.7%) | 207 (38.3%)<sup>a</sup> | 533 (59.3%) | 371 (44.6%)<sup>b</sup> |
| Race: Non-Hispanic white | 347 (58.0%) | 264 (48.8%) | 718 (79.9%) | 554 (66.6%) |
| African American | 251 (42.0%) | 277 (51.2%)<sup>a</sup> | 181 (20.1%) | 278 (33.4%)<sup>b</sup> |
| Pack-years of smoking | 37.4 (20.0) | 36.5 (20.5) | 54.7 (29.3) | 48.5 (26.2)<sup>b</sup> |
| Current smoker | 364 (60.9%) | 337 (62.3%) | 383 (42.6%) | 331 (39.8%) |
| Body mass index, kg/m<sup>2</sup> | 29.0 (5.7) | 30.0 (6.6)<sup>a</sup> | 27.7 (5.9) | 28.9 (6.6)<sup>b</sup> |
| FEV<sub>1</sub> % predicted, post-bronchodilator | 98.2 (11.7) | 95.9 (10.8)<sup>a</sup> | 53.7 (17.6) | 49.0 (17.7)<sup>b</sup> |
| FEV<sub>1</sub>/FVC, post-bronchodilator | 0.79 (0.05) | 0.78 (0.05)<sup>a</sup> | 0.51 (0.13) | 0.50 (0.13)<sup>b</sup> |
| Post-bronchodilator change in FEV<sub>1</sub>, L | 0.09 (0.16) | 0.11 (0.17)<sup>a</sup> | 0.10 (0.17) | 0.11 (0.16) |
| Number of exacerbations in the past year | 0.1 (0.4) | 0.4 (0.9)<sup>a</sup> | 0.5 (1.0) | 1.1 (1.5)<sup>b</sup> |
| Severe exacerbation in past year | 9 (1.5%) | 74 (13.7%)<sup>a</sup> | 145 (16.1%) | 268 (32.2%)<sup>b</sup> |
| Chronic bronchitis | 75 (12.5%) | 109 (20.2%)<sup>a</sup> | 219 (24.4%) | 274 (32.9%)<sup>b</sup> |
| Hay fever | 136 (22.7%) | 271 (50.1%)<sup>a</sup> | 204 (22.7%) | 383 (46.0%)<sup>b</sup> |
| % emphysema (-950HU) | 1.9 (2.4) | 1.7 (2.5) | 12.8 (12.4) | 11.4 (11.6)<sup>b</sup> |
| % gas trapping on expiratory CT scan (-856HU) | 10.0 (8.3) | 9.2 (8.2) | 36.8 (20.2) | 37.0 (19.9) |
| Wall area % of segmental airways | 46.8 (7.3) | 49.4 (7.7)<sup>a</sup> | 54.6 (7.7) | 56.5 (7.8)<sup>b</sup> |
| Total IgE, log10 transformed (SD) | 1.68 (0.67) | 1.87 (0.68)<sup>a</sup> | 1.65 (0.73) | 1.84 (0.72)<sup>b</sup> |
| Total IgE >100 IU/ml | 186 (31.1%) | 231 (42.7%)<sup>a</sup> | 264 (29.4%) | 330 (39.7%)<sup>b</sup> |
| At least 1 positive specific IgE | 181 (30.3%) | 270 (49.9%)<sup>a</sup> | 220 (24.5%) | 362 (43.5%)<sup>b</sup> |

<sup>a</sup>P<0.05 for comparison with no COPD, no asthma
<sup>b</sup>P<0.05 for comparison with COPD, no asthma
<sup>c</sup>Two subjects had failed total IgE assays

In each of the four subgroups based on COPD and asthma, we tested for concordance between having at least one positive specific IgE and an elevated total IgE, using two thresholds (Supplemental Tables 2, 3). Overall, there was better concordance in the subjects with asthma, with or without ACO, and there was better concordance using the higher total IgE cutoff of 100 IU/ml. Based on these results, this threshold was used for subsequent analyses. Due to the potential effects of smoking on IgE levels,^18^ we examined the concordance between elevated total IgE and positive specific IgE stratified by current vs. former smokers (Supplemental Tables 4, 5). A higher proportion of current smokers had both elevated total IgE and at least one positive specific IgE.

### Asthma-COPD overlap

We then examined the various definitions of ACO. Supplemental Table 6 shows the clinical and imaging characteristics of subjects with the strict ACO definition, based on self-report of doctor’s diagnosis of asthma before age 40. The results are generally similar to those for the broad ACO definition based on any self-report of asthma (Table 1) and similar to those that we have previously reported in the entire COPDGene Study.^7^

Of the 831 subjects with the broad definition of ACO, 450 were atopic based on total IgE >100 IU/ml and/or at least one positive specific IgE (Table 2). Compared to subjects with COPD without asthma and without atopy, the subjects with atopic ACO were younger and more commonly African American, though there was no longer a gender difference. These subjects had lower FEV_1_ and more exacerbations. They had a higher prevalence of chronic bronchitis symptoms, and more than half reported a history of hay fever. Chest CT scans showed less emphysema and more airway thickening.

**Table 2:** Subjects with “broad” asthma-COPD overlap (ACO) and atopy. Broad ACO is defined by defined by FEV_1_/FVC<0.7, FEV_1_<80% predicted, and self-report of asthma. Atopy is defined by total IgE>100 IU/ml or at least one positive specific IgE. Mean (SD) or N (%) is shown.

|  | 1. Broad ACO<br>with atopy | 2. COPD, no asthma,<br>no atopy | p-value<br>1 vs 2 | 3. ACO without<br>atopy | p-value<br>1 vs 3 |
| --- | --- | --- | --- | --- | --- |
| N | 450 | 562 |  | 381 |  |
| Age | 61.3 (8.9) | 64.7 (8.3) | <0.001 | 62.4 (8.2) | 0.06 |
| Male gender | 223 (49.6%) | 299 (53.2%) | 0.3 | 147 (38.6%) | 0.002 |
| Race: Non-Hispanic white | 265 (58.9%) | 486 (86.5%) | <0.001 | 288 (75.6%) | <0.001 |
| African American | 185 (41.1%) | 76 (13.5%) |  | 93 (24.4%) |  |
| Pack-years of smoking | 46.5 (26.2) | 55.5 (28.0) | <0.001 | 50.9 (26.1) | 0.02 |
| Current smoker | 195 (43.3%) | 220 (39.2%) | 0.2 | 136 (35.7%) | 0.03 |
| Body mass index, kg/m<sup>2</sup> | 28.9 (6.7) | 27.5 (5.9) | <0.001 | 28.9 (6.5) | 1.0 |
| FEV<sub>1</sub> % predicted, post-bronchodilator | 50.6 (17.1) | 52.7 (17.7) | 0.05 | 47.1 (18.2) | 0.005 |
| FEV<sub>1</sub>/FVC, post-bronchodilator | 0.51 (0.12) | 0.50 (0.13) | 0.5 | 0.48 (0.13) | 0.005 |
| Post-bronchodilator change in FEV<sub>1</sub>, L | 0.11 (0.18) | 0.10 (0.15) | 0.1 | 0.10 (0.15) | 0.4 |
| Number of exacerbations in the past year | 1.1 (1.5) | 0.5 (1.1) | <0.001 | 1.2 (1.4) | 0.3 |
| Severe exacerbation in past year | 145 (32.2%) | 89 (15.8%) | <0.001 | 123 (32.3%) | 1.0 |
| Chronic bronchitis | 148 (32.9%) | 133 (23.7%) | 0.001 | 126 (33.1%) | 1.0 |
| Hay fever | 234 (52%) | 108 (19.2%) | <0.001 | 148 (38.9%) | <0.001 |
| % emphysema (-950HU) | 10.1 (10.6) | 13.6 (13.0) | <0.001 | 12.9 (12.6) | 0.001 |
| % gas trapping on expiratory CT scan (-856HU) | 35.3 (19.9) | 38.0 (20.3) | 0.06 | 38.9 (19.9) | 0.02 |
| Wall area % of segmental airways | 56.6 (7.6) | 53.9 (7.7) | <0.001 | 56.4 (8.1) | 0.7 |

We then examined the 381 subjects with ACO without atopy. Compared to atopic ACO (Table 2), these subjects were more commonly women and non-Hispanic white. They had a greater lifetime smoking intensity but fewer were current smokers. They had lower FEV_1_, but exacerbation rates were not different. This group had a lower prevalence of hay fever compared to atopic ACO, but higher than COPD without asthma or atopy. Compared to atopic ACO, non-atopic ACO had more emphysema, but no difference in airway wall thickening. When subjects with strict ACO definition were subcategorized by atopy, the results were generally similar to broad ACO with atopy (Table 3).

**Table 3:** Subjects with “strict” asthma-COPD overlap (ACO) and atopy. Strict asthma-COPD overlap is defined by FEV_1_/FVC<0.7, FEV_1_<80% predicted, and self-report of a doctor’s diagnosis of asthma before age 40. Atopy is defined by total IgE>100 IU/ml or at least one positive specific IgE. Mean (SD) or N (%) is shown.

|  | 1. Strict ACO with atopy | 2. COPD, no asthma, no atopy | p-value 1 vs 2 | 3. Strict ACO without atopy | p-value 1 vs 3 |
| --- | --- | --- | --- | --- | --- |
| N | 276 | 562 |  | 164 |  |
| Age | 60.0 (9.1) | 64.7 (8.3) | <0.001 | 60.8 (8.1) | 0.3 |
| Male gender | 151 (54.7%) | 299 (53.2%) | 0.7 | 59 (36.0%) | <0.001 |
| Race: Non-Hispanic white | 154 (55.8%) | 486 (86.5%) | <0.001 | 124 (75.6%) | <0.001 |
| African American | 122 (44.2%) | 76 (13.5%) |  | 40 (24.4%) |  |
| Pack-years of smoking | 46.0 (25.4) | 55.5 (28.0) | <0.001 | 47.0 (23.9) | 0.7 |
| Current smoker | 128 (46.4%) | 220 (39.2%) | 0.05 | 63 (38.4%) | 0.1 |
| Body mass index, kg/m<sup>2</sup> | 29.0 (6.9) | 27.5 (5.9) | 0.002 | 29.2 (6.9) | 0.8 |
| FEV<sub>1</sub> % predicted, post-bronchodilator | 51.7 (17.0) | 52.7 (17.7) | 0.4 | 48.1 (19.0) | 0.05 |
| FEV<sub>1</sub>/FVC, post-bronchodilator | 0.51 (0.13) | 0.50 (0.13) | 0.2 | 0.50 (0.14) | 0.3 |
| Post-bronchodilator change in FEV<sub>1</sub>, L | 0.11 (0.18) | 0.10 (0.15) | 0.3 | 0.10 (0.14) | 0.6 |
| Number of exacerbations in the past year | 1.0 (1.5) | 0.5 (1.1) | <0.001 | 1.4 (1.5) | 0.03 |
| Severe exacerbation in past year | 88 (31.9%) | 89 (15.8%) | <0.001 | 58 (35.4%) | 0.5 |
| Chronic bronchitis | 99 (35.9%) | 133 (23.7%) | <0.001 | 54 (32.9%) | 0.6 |
| Hay fever | 156 (56.5%) | 108 (19.2%) | <0.001 | 68 (41.5%) | 0.009 |
| % emphysema (-950HU) | 8.8 (9.9) | 13.6 (13.0) | <0.001 | 10.7 (11.8) | 0.1 |
| % gas trapping on expiratory CT scan (-856HU) | 33.0 (19.6) | 38.0 (20.3) | 0.002 | 36.3 (20.7) | 0.1 |
| Wall area % of segmental airways | 57.2 (7.9) | 53.9 (7.7) | <0.001 | 57.2 (8.0) | 0.9 |

Figure 1 demonstrates the overlaps between the subjects with asthma, COPD, and atopy. There is high overlap between asthma and COPD (N=831 broad definition ACO), which was one of the selection criteria for this COPDGene substudy. As above, a substantial number of subjects with ACO did not have atopy (N=382). Conversely, there are many subjects with atopic COPD, who also have the diagnosis of asthma (N=450). Figure 2 shows similar results, using the strict asthma definition.

**Figure 1:**
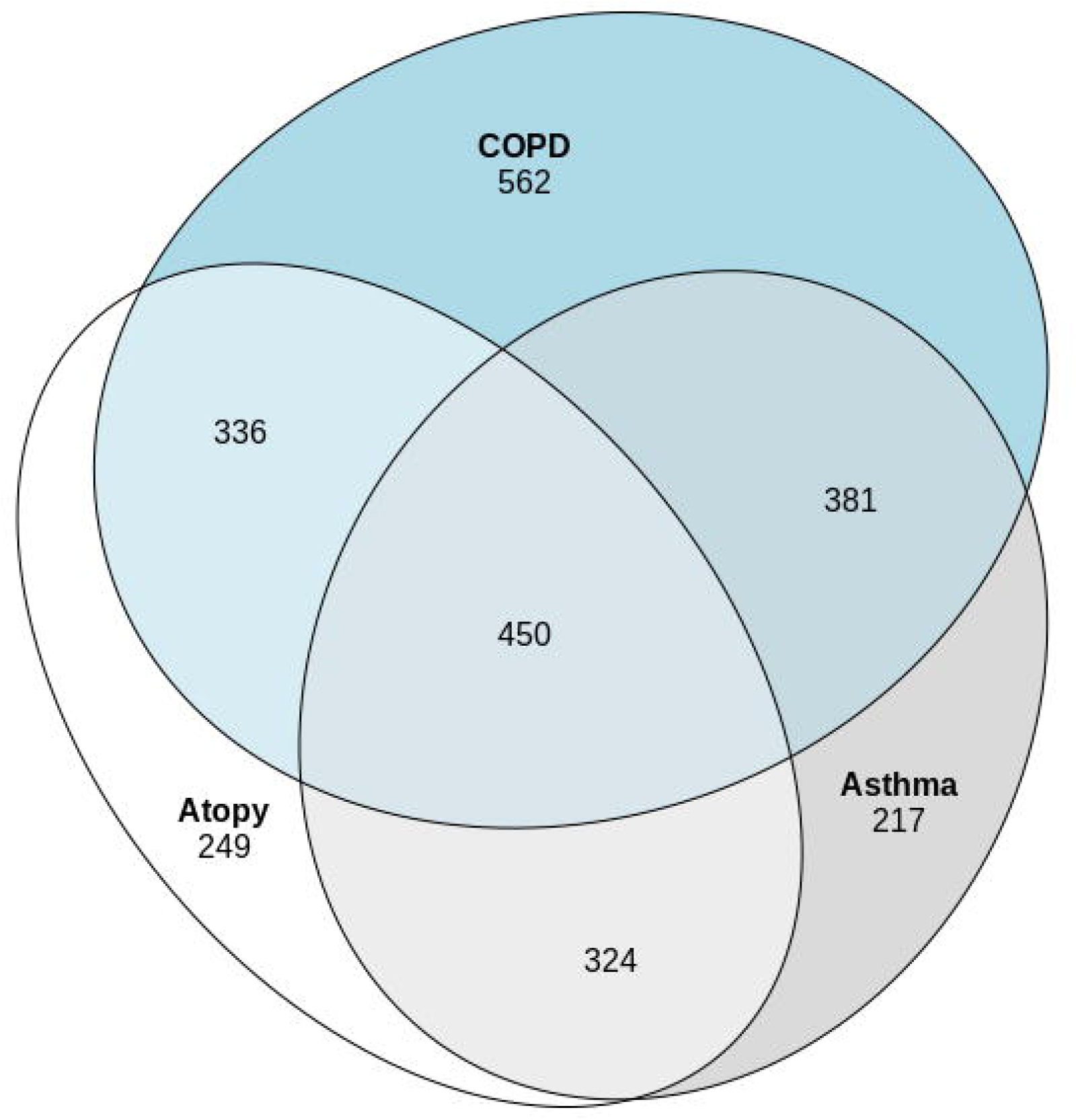
Venn diagram showing overlaps between asthma, COPD, and atopy. Atopy is defined by total IgE >100 IU/ml or at least one positive specific IgE.

**Figure 2:**
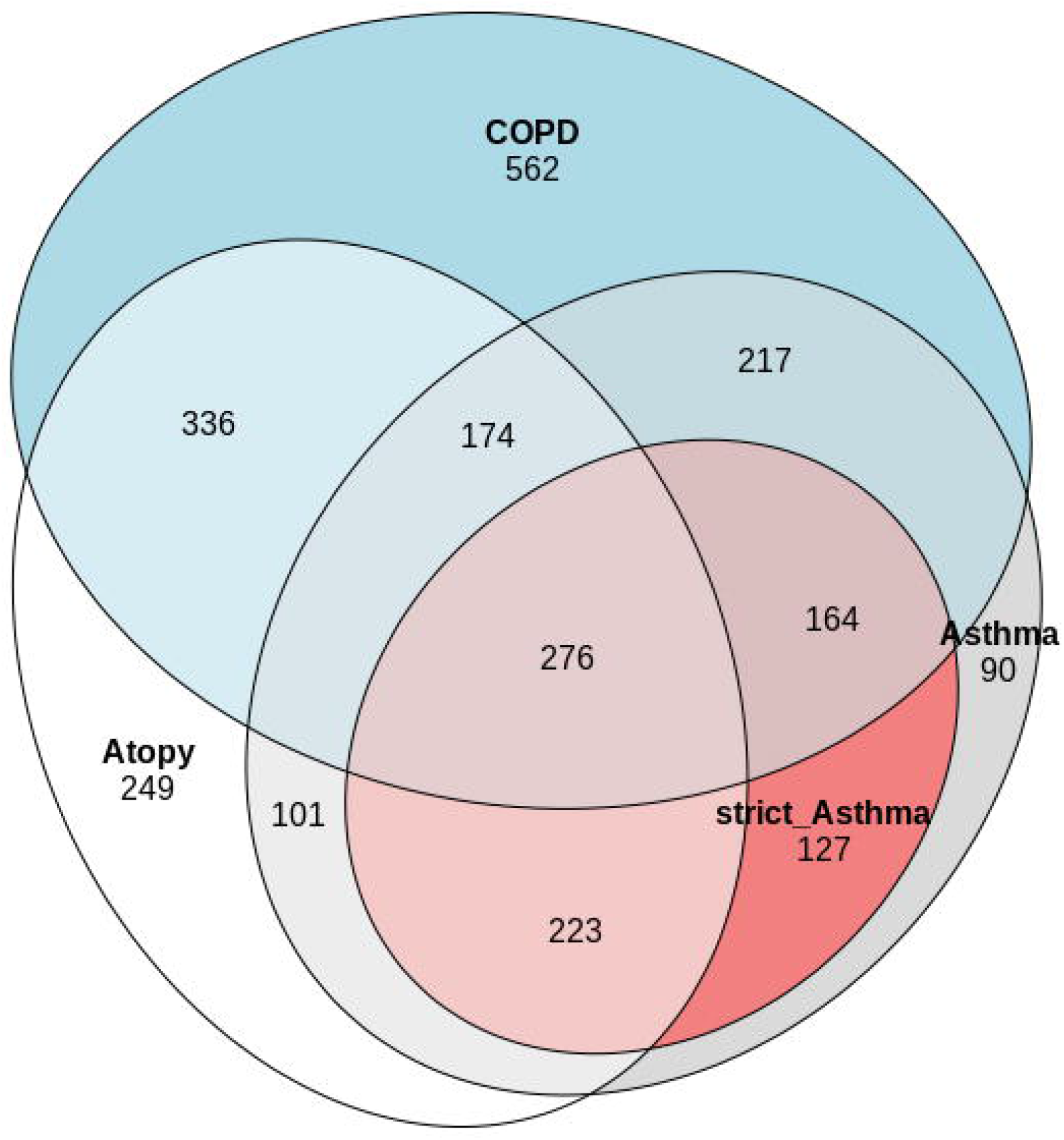
Venn diagram of asthma, COPD, and atopy, including a strict definition of asthma, defined by self-report of a doctor’s diagnosis of asthma before age 40.

### Comparison of ACO definitions

We tested three possible ACO definitions for association with exacerbations: COPD with strict asthma, COPD with asthma and atopy, and COPD with strict asthma and atopy. Each definition was similarly associated with number of exacerbations (Table 4) or the presence of a severe exacerbation (Supplemental Table 7) in the year prior to enrollment, using linear or logistic regression models respectively. Only the first two definitions were significant predictors of exacerbations or severe exacerbations in the longitudinal follow-up (Table 4). All three definitions were similarly associated with airway wall thickening on chest CT scans (Supplemental Table 8).

**Table 4:**
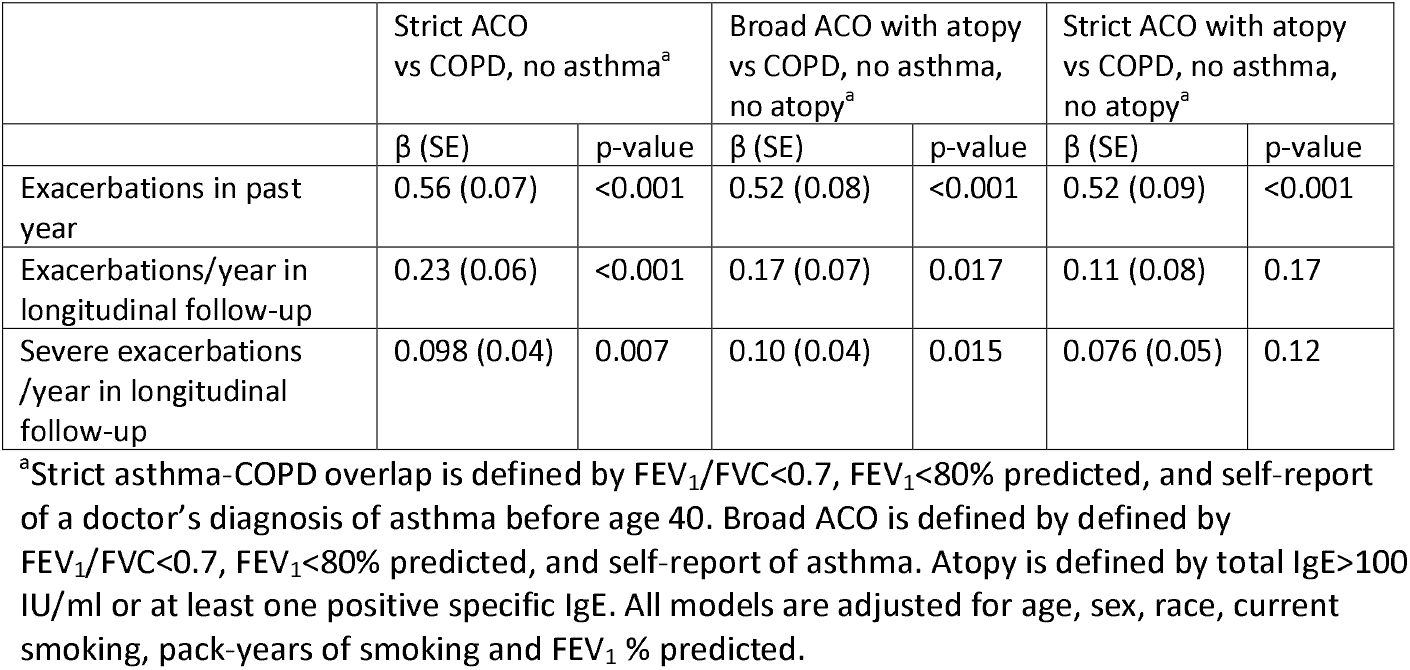
Linear regression models for exacerbations outcomes

## Discussion

In this study, we measured total and specific IgE levels in a large epidemiologic study of smokers with and without COPD, which included subjects with a history of asthma. We used the presence of atopy to refine the definition of ACO, either based on COPD with any asthma history or our previous ACO definition based on self-report of a doctor’s diagnosis of asthma before the age of 40. Similar to our previous results, ACO subjects by any definition were younger, more commonly female or African American, and had similar levels of lung function impairment despite lower lifetime smoking intensity.^6,7^ ACO subjects had more symptoms of hay fever and chronic bronchitis. Despite different subjects meeting each of the ACO criteria, each definition was associated with COPD exacerbations and airway wall thickening on Chest CT scans. IgE levels did not clearly refine the clinical diagnosis of ACO. However, we did identify a subgroup of ACO without atopy, who were more likely to be non-Hispanic white women with higher lifetime smoking intensity but less current smoking, lower lung function, more emphysema but similar airway wall thickening and exacerbation rates.

Previous studies have examined IgE levels in ACO. Soler-Cataluna devised a definition of ACO, based on expert consensus, that included high IgE as a minor criterion, though no specific IgE threshold was provided.^19^ This definition was applied to a observational study of 3125 COPD patients in Spain.^20^ As expected, subjects with ACO had higher IgE levels compared to other COPD patients. A Japanese study compared 37 ACO patients, defined based on GINA/GOLD criteria,^5^ to 220 usual COPD, finding ACO subjects had a higher prevalence of elevated total IgE (>173 IU/ml, based on the reference range of the hospital laboratory) or a positive specific IgE to a panel of allergens largely similar to those in the present study.^21^ In a longitudinal study of 831 COPD patients in Spain, Cosio et al. defined ACO by the GINA/GOLD criteria, augmented with major and minor criteria, the latter of which included total IgE >100 IU/ml.^22^ As expected, subjects with ACO had higher IgE levels than no ACO. Almost all subjects with ACO continued to meet criteria at 1 year follow-up and had better survival than non-ACO subjects.

Even when using a strict definition of ACO based on clinical history and spirometry, we found subjects with and without atopy. Asthma is recognized to be a heterogeneous condition, and our study adds evidence to support that ACO is likely to be heterogeneous as well.^1^ In a study of 109 ACO patients, less than half were considered to have Th2-inflammation, based on blood or sputum eosinophilia.^23^ Barnes proposed subdividing ACO into eosinophilic, neutrophilic, and paucigranulocytic subtypes, based on sputum inflammatory cell patterns, and has suggested therapies for each subtype.^24^ Ghebre et al. defined eosinophilic, neutrophilic, and mixed subgroups of asthma and COPD patients, based on sputum cytokines.^25^ Interleukin (IL)-17 is one driver of neutrophilic inflammation in severe asthma and potentially COPD.^26^ COPD subjects with an IL-17 bronchial gene expression pattern may be less responsive to inhaled corticosteroids.^27^

Agusti et al. introduced the concept of treatable traits, which favors the use of treatable phenotypes over specific diagnostic categories of airway disease, such as COPD or asthma.^28^ Elevated total IgE is a treatable trait in asthma, which can be targeted with omalizumab, a monoclocal antibody to IgE. Omalizumab has been shown to be effective in the treatment of ACO in an Australian registry study^29^ and a post-hoc analysis of the PROSPERO clinical trial.^30^ Eosinophilia is another potential treatable trait in COPD, based on studies demonstrating effectiveness of mepolizumab, an anti-IL-5 antibody in COPD subjects with elevated blood eosinophils.^31^ We have previously shown that an eosinophil threshold >300 cells/ul had the strongest association with exacerbations in COPDGene, but had limited overlap with a clinical definition of ACO.^32^ However, the eosinophil counts were measured at the COPDGene Phase 2 (5-year) visit, so we could not directly compare with the IgE levels in this study, which were measured at the baseline visit.

Besides the inability to directly correlate IgE levels with eosinophil counts, there are several limitations to our study. We relied on a self-report of asthma, which could be subject to recall bias or diagnostic misclassification. However, we have used this definition of ACO in previous studies in COPDGene, identifying both clinical and genetic associations.^6,7^ We assayed a limited number of specific IgE tests, based on a previous study in COPD.^10^ These allergens also showed some of the highest sensitization rates in the National Health and Nutrition Examination Survey, a sample of the U.S. population.^33^ Our study focused on indoor aeroallegens, since outdoor allergens may vary more by season and by region in the multi-center study, and since COPD patients usually spend more time indoors.^34^ Perhaps because of the limited panel of specific IgE tests, we found that elevated total IgE captured the majority of the atopic ACO subjects (Supplemental Table 3). In future studies, it would be relevant to examine a larger panel of specific IgE measurements, along with corresponding assessment of allergen exposures, which was not available in COPDGene.

By measuring total and specific IgE in a study of smokers with and without COPD, we identified subjects with atopic asthma-COPD overlap, who were similar to subjects with a clinical definition of ACO. IgE did not improve prediction of exacerbation rates or airway wall thickening on chest CT scans. Total and specific IgE measurements allowed us to subdivide ACO subjects into those with and without atopy; the latter were more likely to be white men with greater lifetime smoking intensity, lower lung function and more emphysema on chest CT scans. It is likely that atopic ACO subjects represent a subtype of COPD with a different mechanism of disease risk and progression, who would benefit from alternative therapeutic considerations. Further investigation using the wealth of molecular data in COPDGene will be required to identify biologic mechanisms underlying non-atopic ACOS. Currently, IgE levels are not a standard test in COPD management, nor are they included in consensus definitions for ACO.^2,4,5^ However, peripheral eosinophilia is increasingly recognized as a treatable trait in COPD.^28,35^ Future studies will be required to determine the overlap between atopic ACO and eosinophilic COPD and whether IgE testing should be considered as part of the evaluation and management of a COPD patient.

## Data Availability

COPDGene Study data is available on dbGAP, accession phs000179.

## Abbreviations

ACO: asthma-COPD overlap
CT: computed tomography
FEV_1_: forced expiratory volume in 1 sec
FVC: forced vital capacity
IgE: immunoglobulin E

## Acknowledgements

COPDGene^®^ Investigators – Core Units

## Administrative Center

James D. Crapo, MD (PI); Edwin K. Silverman, MD, PhD (PI); Barry J. Make, MD; Elizabeth A. Regan, MD, PhD Genetic Analysis Center: Terri Beaty, PhD; Ferdouse Begum, PhD; Peter J. Castaldi, MD, MSc; Michael Cho, MD; Dawn L. DeMeo, MD, MPH; Adel R. Boueiz, MD; Marilyn G. Foreman, MD, MS; Eitan Halper-Stromberg; Lystra P. Hayden, MD, MMSc; Craig P. Hersh, MD, MPH; Jacqueline Hetmanski, MS, MPH; Brian D. Hobbs, MD; John E. Hokanson, MPH, PhD; Nan Laird, PhD; Christoph Lange, PhD; Sharon M. Lutz, PhD; Merry-Lynn McDonald, PhD; Margaret M. Parker, PhD; Dandi Qiao, PhD; Elizabeth A. Regan, MD, PhD; Edwin K. Silverman, MD, PhD; Emily S. Wan, MD; Sungho Won, Ph.D.; Phuwanat Sakornsakolpat, M.D.; Dmitry Prokopenko, Ph.D.

## Imaging Center

Mustafa Al Qaisi, MD; Harvey O. Coxson, PhD; Teresa Gray; MeiLan K. Han, MD, MS; Eric A. Hoffman, PhD; Stephen Humphries, PhD; Francine L. Jacobson, MD, MPH; Philip F. Judy, PhD; Ella A. Kazerooni, MD; Alex Kluiber; David A. Lynch, MB; John D. Newell, Jr., MD; Elizabeth A. Regan, MD, PhD; James C. Ross, PhD; Raul San Jose Estepar, PhD; Joyce Schroeder, MD; Jered Sieren; Douglas Stinson; Berend C. Stoel, PhD; Juerg Tschirren, PhD; Edwin Van Beek, MD, PhD; Bram van Ginneken, PhD; Eva van Rikxoort, PhD; George Washko, MD; Carla G. Wilson, MS;

## PFT QA Center, Salt Lake City, UT

Robert Jensen, PhD

## Data Coordinating Center and Biostatistics, National Jewish Health, Denver, CO

Douglas Everett, PhD; Jim Crooks, PhD; Camille Moore, PhD; Matt Strand, PhD; Carla G. Wilson, MS

## Epidemiology Core, University of Colorado Anschutz Medical Campus, Aurora, CO

John E. Hokanson, MPH, PhD; John Hughes, PhD; Gregory Kinney, MPH, PhD; Sharon M. Lutz, PhD; Katherine Pratte, MSPH; Kendra A. Young, PhD

## Mortality Adjudication Core

Surya Bhatt, MD; Jessica Bon, MD; MeiLan K. Han, MD, MS; Barry Make, MD; Carlos Martinez, MD, MS; Susan Murray, ScD; Elizabeth Regan, MD; Xavier Soler, MD; Carla G. Wilson, MS

## Biomarker Core

Russell P. Bowler, MD, PhD; Katerina Kechris, PhD; Farnoush Banaei-Kashani, Ph.D

COPDGene^®^ Investigators – Clinical Centers

## Ann Arbor VA

Jeffrey L. Curtis, MD; Carlos H. Martinez, MD, MPH; Perry G. Pernicano, MD

## Baylor College of Medicine, Houston, TX

Nicola Hanania, MD, MS; Philip Alapat, MD; Mustafa Atik, MD; Venkata Bandi, MD; Aladin Boriek, PhD; Kalpatha Guntupalli, MD; Elizabeth Guy, MD; Arun Nachiappan, MD; Amit Parulekar, MD;

## Brigham and Women’s Hospital, Boston, MA

Dawn L. DeMeo, MD, MPH; Craig Hersh, MD, MPH; Francine L. Jacobson, MD, MPH; George Washko, MD

## Columbia University, New York, NY

R. Graham Barr, MD, DrPH; John Austin, MD; Belinda D’Souza, MD; Gregory D.N. Pearson, MD; Anna Rozenshtein, MD, MPH, FACR; Byron Thomashow, MD

## Duke University Medical Center, Durham, NC

Neil MacIntyre, Jr., MD; H. Page McAdams, MD; Lacey Washington, MD

## HealthPartners Research Institute, Minneapolis, MN

Charlene McEvoy, MD, MPH; Joseph Tashjian, MD

## Johns Hopkins University, Baltimore, MD

Robert Wise, MD; Robert Brown, MD; Nadia N. Hansel, MD, MPH; Karen Horton, MD; Allison Lambert, MD, MHS; Nirupama Putcha, MD, MHS

## Los Angeles Biomedical Research Institute at Harbor UCLA Medical Center, Torrance, CA

Richard Casaburi, PhD, MD; Alessandra Adami, PhD; Matthew Budoff, MD; Hans Fischer, MD; Janos Porszasz, MD, PhD; Harry Rossiter, PhD; William Stringer, MD

## Michael E. DeBakey VAMC, Houston, TX

Amir Sharafkhaneh, MD, PhD; Charlie Lan, DO

## Minneapolis VA

Christine Wendt, MD; Brian Bell, MD

## Morehouse School of Medicine, Atlanta, GA

Marilyn G. Foreman, MD, MS; Eugene Berkowitz, MD, PhD; Gloria Westney, MD, MS

## National Jewish Health, Denver, CO

Russell Bowler, MD, PhD; David A. Lynch, MB

## Reliant Medical Group, Worcester, MA

Richard Rosiello, MD; David Pace, MD

## Temple University, Philadelphia, PA

Gerard Criner, MD; David Ciccolella, MD; Francis Cordova, MD; Chandra Dass, MD; Gilbert D’Alonzo, DO; Parag Desai, MD; Michael Jacobs, PharmD; Steven Kelsen, MD, PhD; Victor Kim, MD; A. James Mamary, MD; Nathaniel Marchetti, DO; Aditi Satti, MD; Kartik Shenoy, MD; Robert M. Steiner, MD; Alex Swift, MD; Irene Swift, MD; Maria Elena Vega-Sanchez, MD

## University of Alabama, Birmingham, AL

Mark Dransfield, MD; William Bailey, MD; Surya Bhatt, MD; Anand Iyer, MD; Hrudaya Nath, MD; J. Michael Wells, MD

## University of California, San Diego, CA

Joe Ramsdell, MD; Paul Friedman, MD; Xavier Soler, MD, PhD; Andrew Yen, MD

## University of Iowa, Iowa City, IA

Alejandro P. Comellas, MD; Karin F. Hoth, PhD; John Newell, Jr., MD; Brad Thompson, MD

## University of Michigan, Ann Arbor, MI

MeiLan K. Han, MD, MS; Ella Kazerooni, MD; Carlos H. Martinez, MD, MPH

## University of Minnesota, Minneapolis, MN

Joanne Billings, MD; Abbie Begnaud, MD; Tadashi Allen, MD

## University of Pittsburgh, Pittsburgh, PA

Frank Sciurba, MD; Jessica Bon, MD; Divay Chandra, MD, MSc; Carl Fuhrman, MD; Joel Weissfeld, MD, MPH

## University of Texas Health Science Center at San Antonio, San Antonio, TX

Antonio Anzueto, MD; Sandra Adams, MD; Diego Maselli-Caceres, MD; Mario E. Ruiz, MD

## Author contributions

Study design and data collection: CPH, LPH, NP

Data analysis: CPH, S. Zacharia, RPAC, AM, S. Zarei

Manuscript writing and editing: All authors

